# A systematic review of the knowledge, attitudes, and practices of physicians, health workers, and the general population about Coronavirus disease 2019 (COVID-19)

**DOI:** 10.1101/2020.10.04.20206094

**Authors:** Saeede Saadatjoo, Maryam Miri, Soheil Hassanipour, Hosein Ameri, Morteza Arab-Zozani

## Abstract

**Background:** Understanding people, physicians, and healthcare workers’ knowledge, attitude, and practices (KAPs) can help to achieve the outcomes of planned behavior. The aim of this study was to investigate and synthesize the current evidence on KAPs regarding COVID-19.

**Methods:** We conducted a systematic search on PubMed/LitCovid, Scopus, and Web of Sciences databases for papers in the English language only, up to 1 Jul 2020. We used the Joanna Briggs Institute (JBI) checklist developed for cross-sectional studies to appraise the quality of the included studies. All stages of the review conducted by two independent reviewers and potential discrepancies solved with a consultation with a third reviewer. We reported the result as number and percentage. PROSPERO registration code: (CRD42020186755).

**Results:** Fifty-two studies encompassing 49786 participants were included in this review. 45.76% of the participants were male. The mean age of the participants was 32.6 years. 44.2% of the included studies were scored as good quality, 46.2% as fair quality, and remaining (9.6%) as low quality. 30.76% examined all three components of the KAPs model. The knowledge component was reported as good, fair, and poor in 59%, 34%, and 7%, respectively. Of the studies that examined the attitude component, 82% reported a positive attitude, 11% a fairly positive attitude, and 7% a negative attitude. For the practice component, 52% reported good practice, 44% fair practice, and 4% poor practice.

**Conclusion:** This systematic review showed that the overall KAP components in the included studies were at an acceptable level. In general, knowledge was at a good level, the attitude was positive and practice was at a fairly good level. Using an integrated international system can help better evaluate these components and compare them between countries.

## Introduction

Coronavirus disease 2019 (COVID-19) was reported on 31st December 2019 from Wuhan, China, and announced by the World Health Organization (WHO) as a pandemic on 11th March 2020 [1, 2]. To date, it was estimated that about 30 million people were infected with COVID-19 worldwide, of which about one million have died [3].

COVID-19 is characterized by a number of flu-like symptoms including fever, respiratory problems (dry cough, shortness of breath or difficulty breathing, sore throat), chills, headache, and loss of taste. Also, this disease is much more severe with men, higher age groups, and patients with other pre-existing conditions, such as cardiovascular disease, chronic respiratory disease, diabetes, and hypertension [4, 5]. Based on existing evidence, about 81% of COVID-19 cased is mild, 14% are severe and 5 % is critical. The median time from symptoms onset to clinical recovery is approximately two weeks for mild cases and three to six weeks for severe or critical cases [6]. The incubation period for this disease was reported as 2-14 days based on WHO reports. The mortality rate for this disease if different among countries and was reported between two% and 5% [7, 8]. The most important ways to prevent this disease are to use a mask and maintain social distance [9-11]. So far, there have been several cases of infection in the general public, especially doctors and medical staff, some of which have led to death [12-14].

Considering the extent and progress of COVID-19 disease and it’s major effects on economic, social, political, and cultural dimensions of all countries [15, 16], it is essential that people with COVID-19 are provoked, informed, and engaged in all aspect of the disease. From the onset of the disease until now, various studies conducted worldwide have investigated on this disease and some of these studies have examined the knowledge, attitudes, and practices (KAPs) of people with COVID-19. Having enough knowledge about a disease can always affect people’s attitudes and practices, and on the other hand, improper attitudes and practices can increase the risk of disease and death. Therefore, understanding people, physicians, and healthcare workers’ KAPs and knowing potential risk factors can help to achieve the outcomes of planned behavior [17, 18].

Given the importance of the issue, conducting a review of studies that have examined the KAPs of individuals and summarizing the results can provide solid evidence for decision-makers in all countries to better manage the disease. Thus, this study aimed at conducting a systematic review to synthesize current evidence on KAPs of people with COVID-19 worldwide.

## Materials and Methods

### Protocol and registration

We conducted a systematic review of the existing evidence related to KAPs of COVID-19 patients worldwide following Preferred Reporting Items for Systematic Reviews and Meta-analyses (PRISMA) statements (Appendix Supplementary file 1) [19]. We also registered a protocol for this systematic review in the International Prospective Register of Systematic Reviews [20] (CRD42020186755).

### Eligibility criteria

We included all studies which met the following inclusion criteria: 1) cross-sectional survey; 2) investigate at least one component of the KAPs model regarding COVID-19 patients worldwide; 3) published, in-press or preprint original paper; 4) in English; 5) with a sample of the general population, physicians or other healthcare workers. No restrictions were applied to the setting, time, or quality of the study.

### Information sources, search and study selection

We search the PubMed/LitCovid, Scopus, and Web of Sciences for papers in the English language only, up to 1 Jul 2020 (MA-Z). We also conducted a search in Google Scholar for retrieving studies that were not cited in the above-mentioned databases. In addition, the reference lists to final articles were hand-searched. The keywords used in the search were: attitude, knowledge, practice, awareness, perception, action, COVID-19, coronavirus disease, SARS-CoV-2, and severe acute respiratory syndrome coronavirus 2. The full search strategy for the Scopus database is provided in Appendix Supplementary file 2. When the search was complete, all records were transferred to the Endnote software (V. X8; Clarivate Analytics, Philadelphia, PA) and duplicates were removed. Then, studies based on the title, abstract, and full text were screened by two researchers independently by considering the pre-specified eligibility criteria (MM and SS). Disagreements were solved through consultation with a third researcher (MA-Z).

### Data collection process and data item

Two researchers independently engaged in the data collection process and extracted data including author, year, journal name, location, study design, data collection tools, sample size, focusing group, mean age or range, gender percent, and result related to KAPs model components (MM and SS). Potential disagreements were solved through consultation with a third researcher (MA-Z).

### Quality appraisal

Included studies were critically appraised by two researchers independently (MM and SS). We used the Joanna Briggs Institute (JBI) checklist developed for cross-sectional studies to appraise the quality of the included studies [21]. This checklist contains eight simple and clear questions that cover topics such as inclusion criteria for sample; details about study subjects and setting; validity and reliability; criteria for measurement of the condition; confounding variables; and statistical analysis [22]. The answer to each questions is yes, no, unclear, and not applicable. Potential discrepancies were resolved by consultation with a third researcher (SH).

### Synthesis of results

Due to potential heterogeneity between included studies the meta-analysis was not conducted. Therefore we carried out the descriptive analysis in most sections and report the pooled data as number or percentage for similar data items. We used Microsoft Excel software to design the charts. Also, we report the result of the included studies in a narrative manner.

## Results

### Study selection

A total of 4829 records were retrieved from our database search. After removing duplicate, 3085 records were screened by title, abstract, and full-text based on eligibility criteria, of which fifty-two studies were included in the final review [23-74]. The PRISMA flow diagram for the complete study selection process is presented in Fig 1.

**Figure 1:**
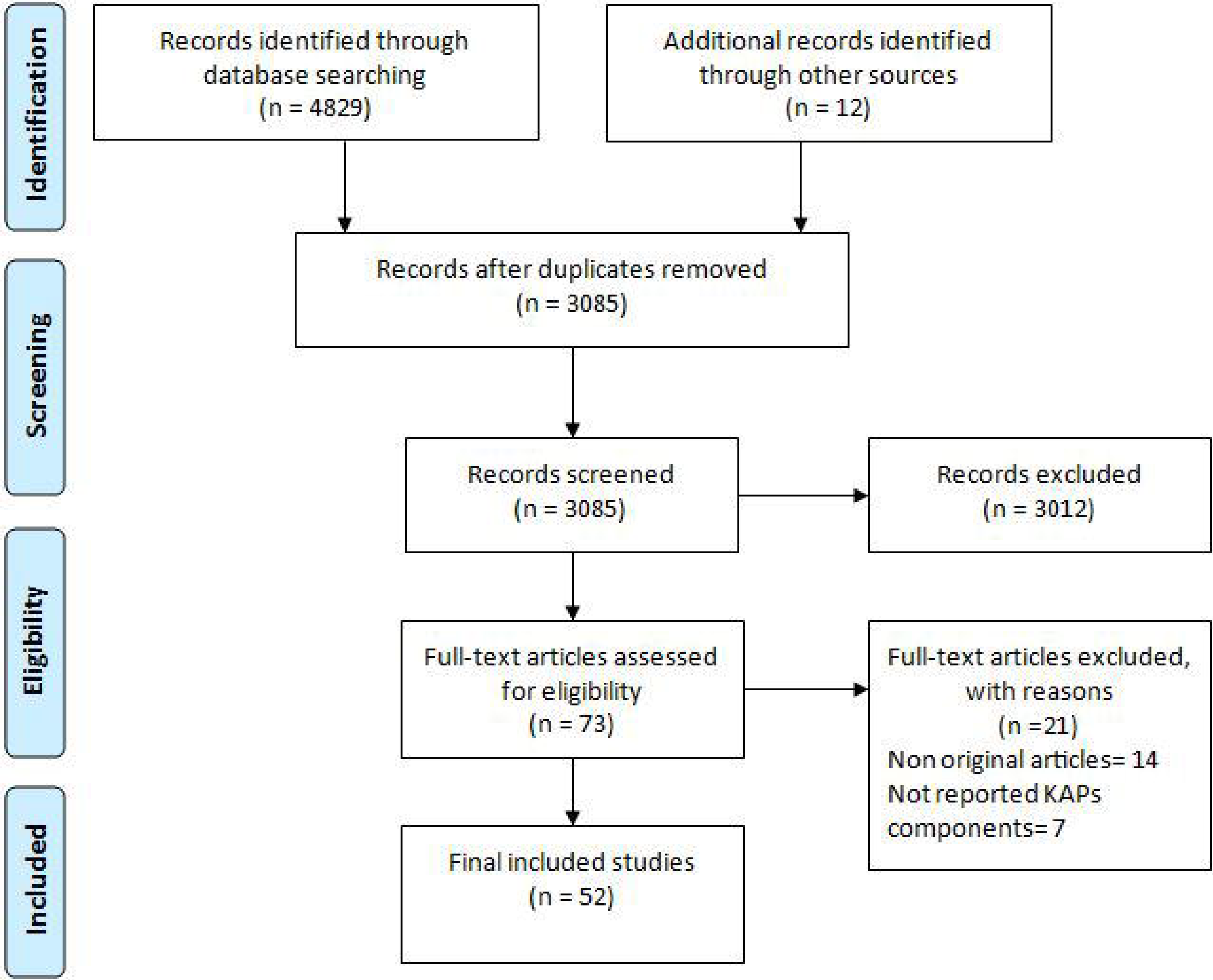
PRISMA flow diagram.

### Study characteristics

Fifty-two studies encompassing 49786 participants were included. Also, 45.76% of the participants were male. The mean age of the participants was 32.6 years. Most studies were from Asia, Africa, South America, Europe, and multinational, respectively (Fig 2A). The most important method of data collection was online questionnaires (Fig 2B). Focusing groups were diverse between the studies and included health care workers, students, general population, adult population, patients groups, residents, and pregnant women (Fig 2C). Most studies examined all three components of the KAPs model, but some studies examined two components or one component. More details about the characteristics of included studies are presented in Table 1.

**Table 1:**
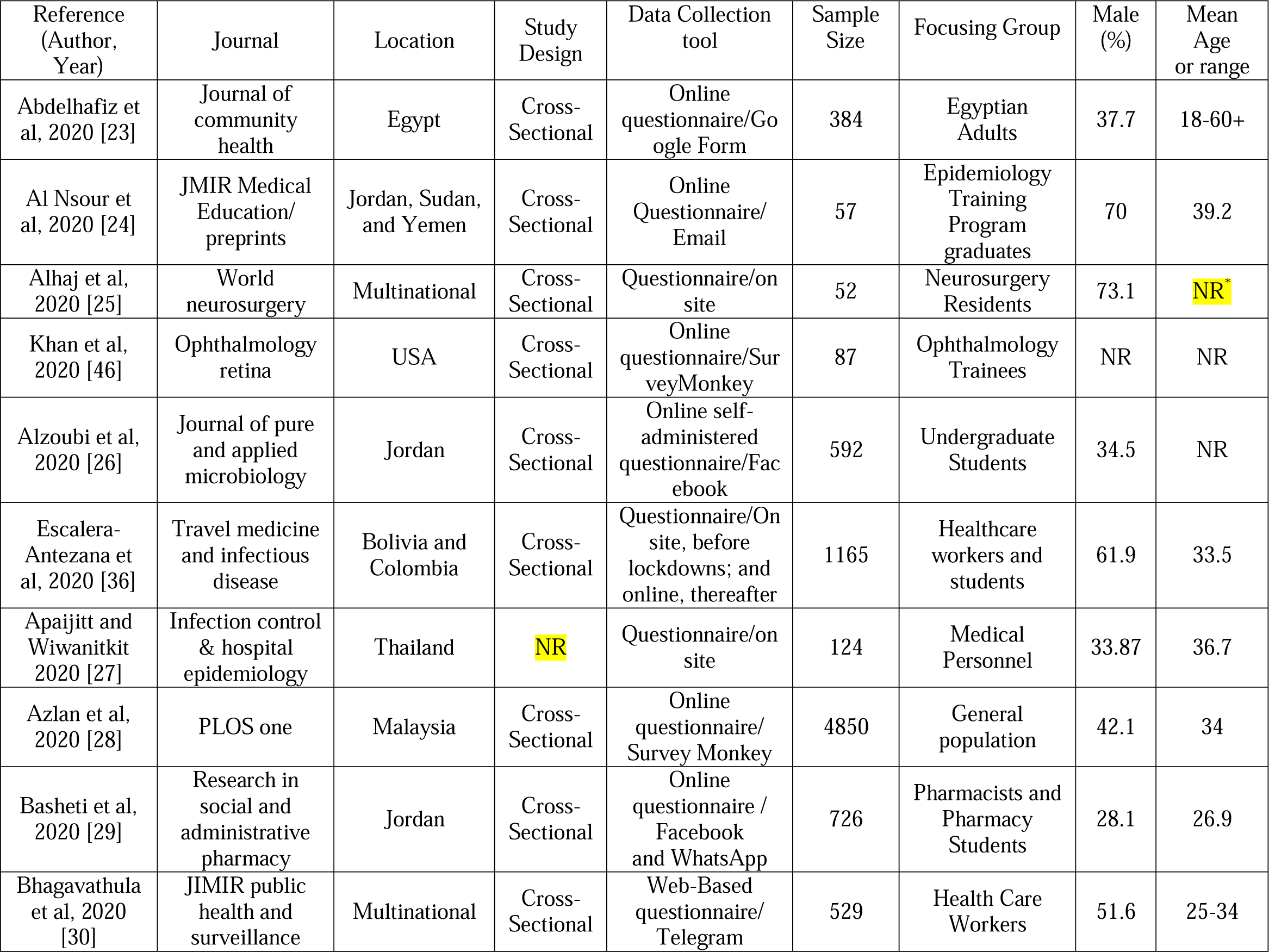
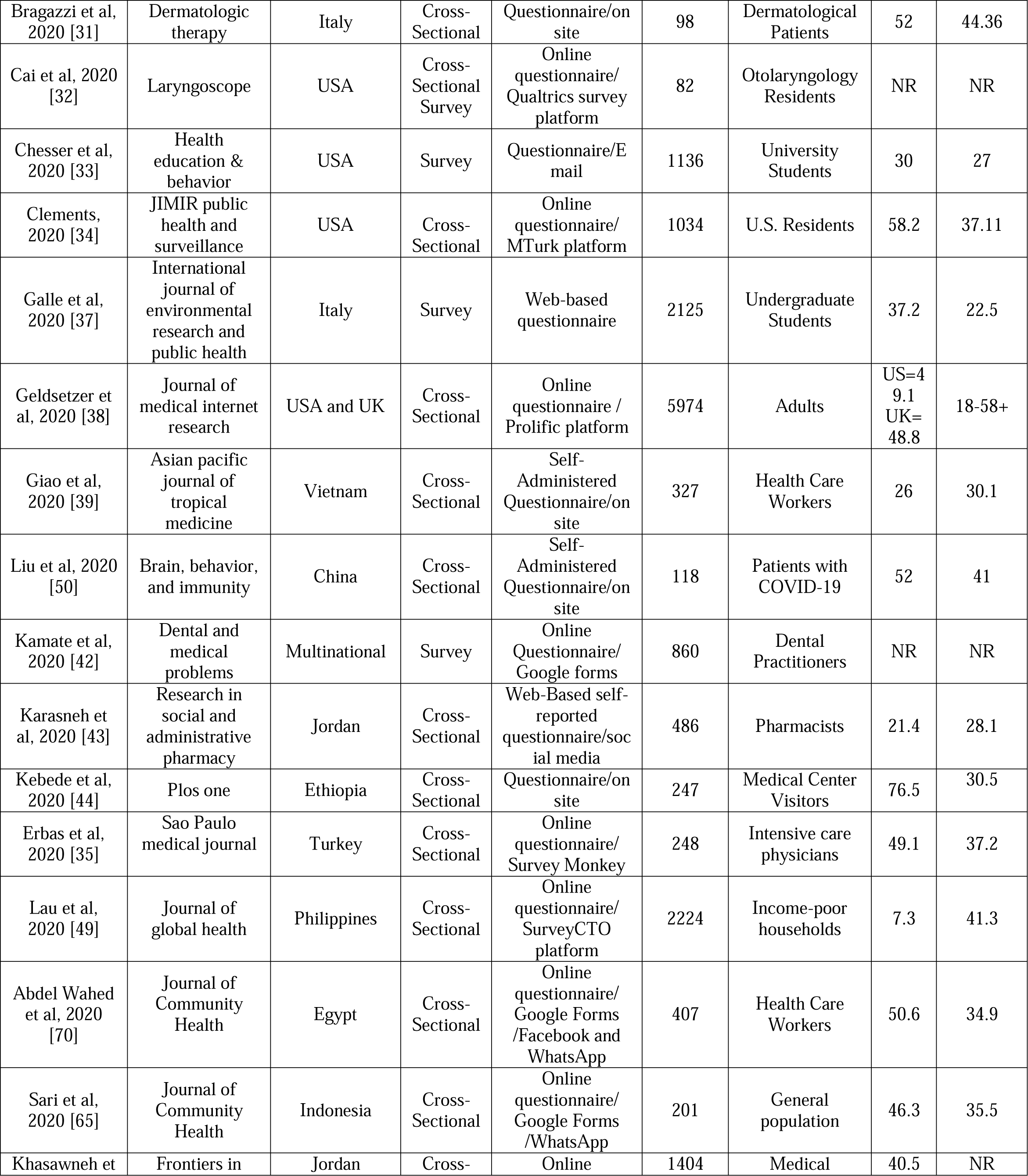
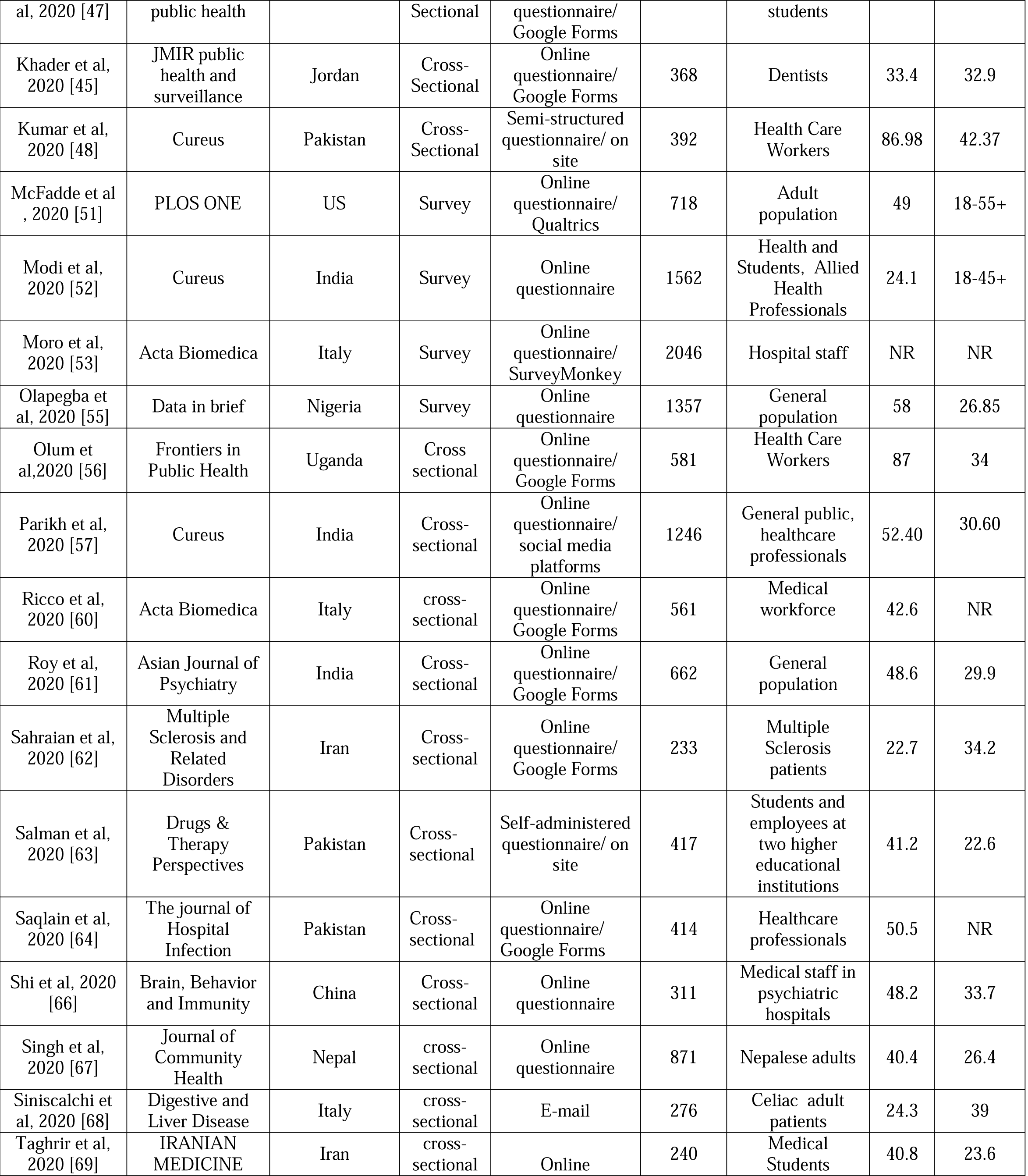
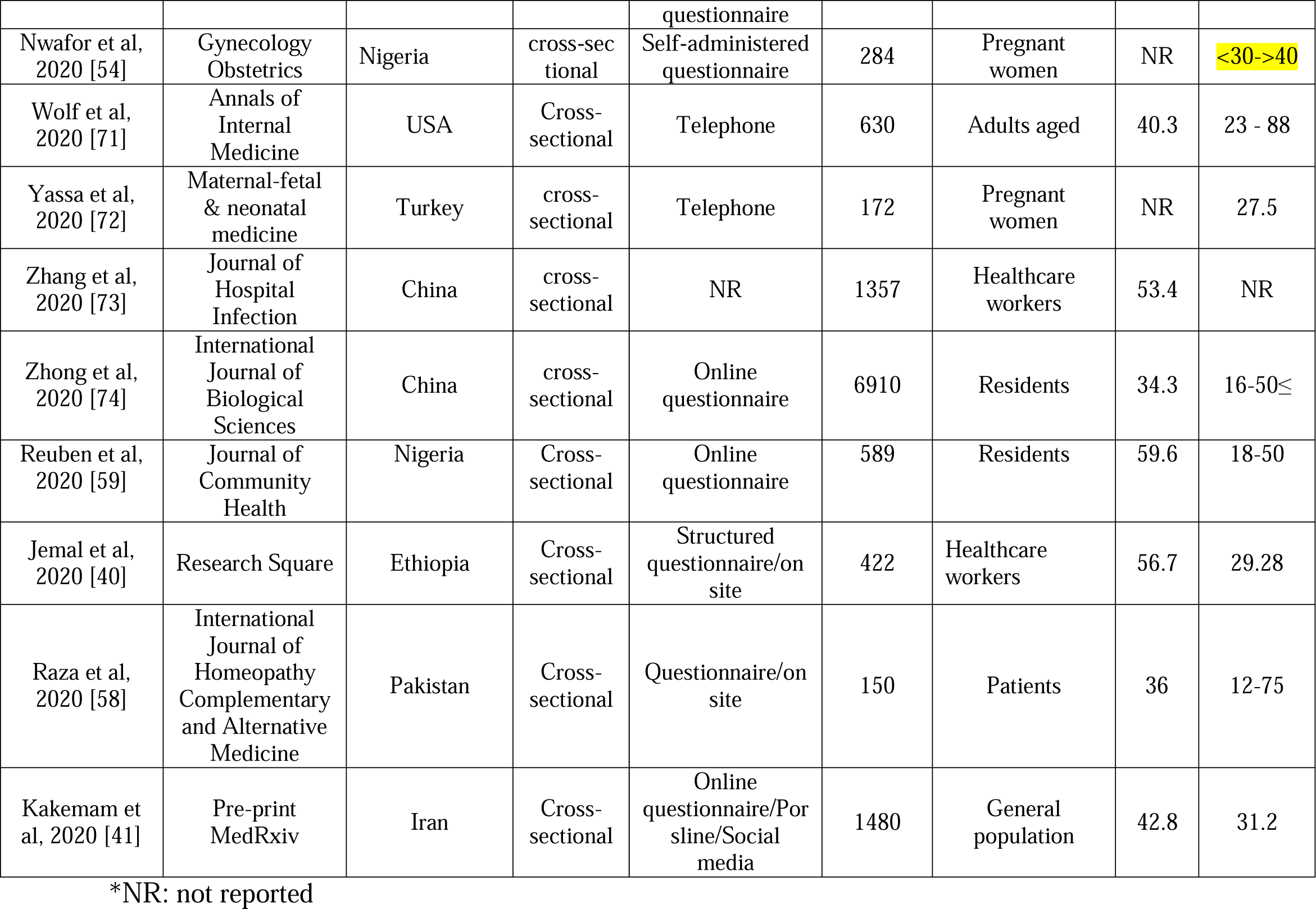
Summary characteristics of the included studies.

**Figure 2:**
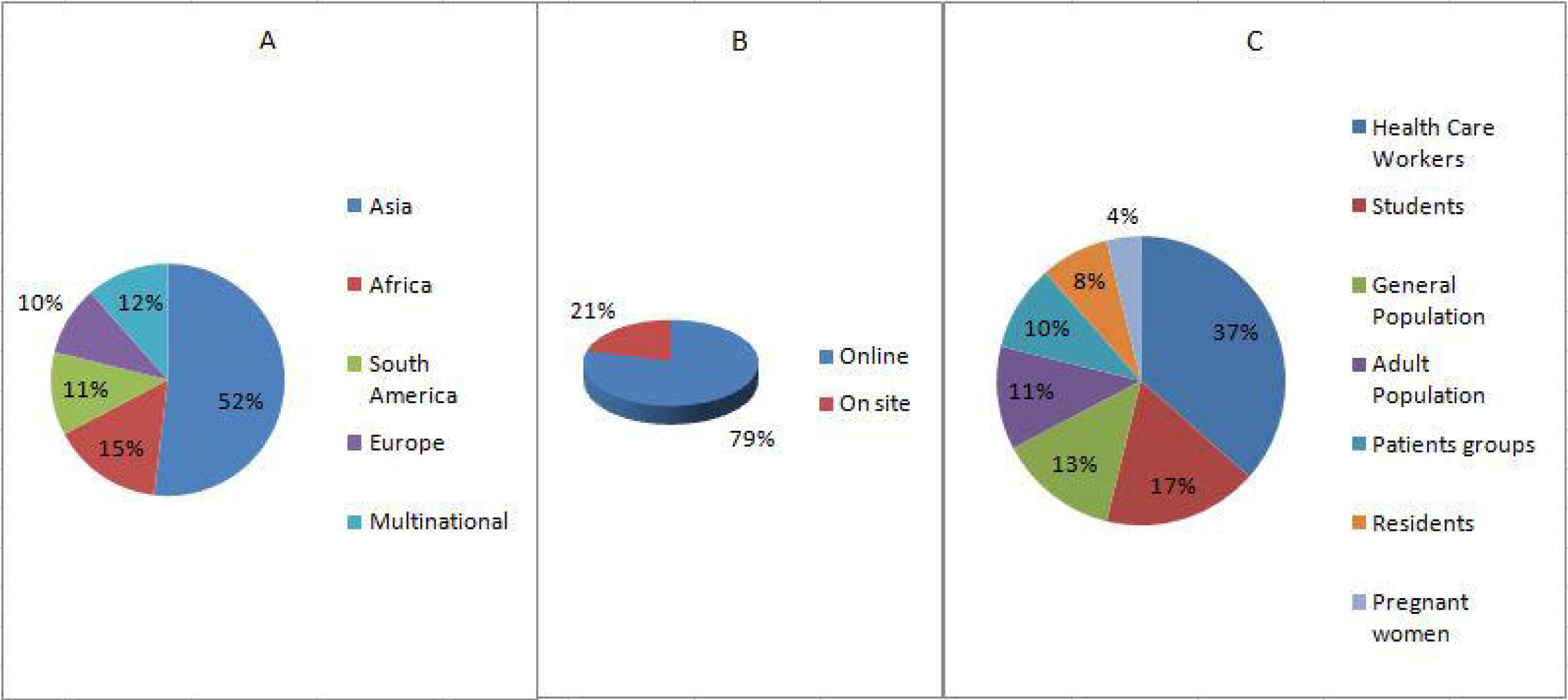
The percentage of the included studies based on location (A), data collection methods (B), and focusing group (C).

### Quality appraisal

The overall mean quality score of the included studies was 4.90. Of the included studies, 23 studies (44.2%) were scored as good quality (score ≥6), 24 (46.2%) as fair quality (score 3-5), and remaining (9.6%) as low quality (score <3) (Fig 3). The lowest and highest quality scores in the studies were two and six, respectively. None of the studies scored on questions 5 and 6, which were related to identification and deal with confounding variables in the studies (for more details about items see Appendix Supplementary file 3.

**Figure 3:**
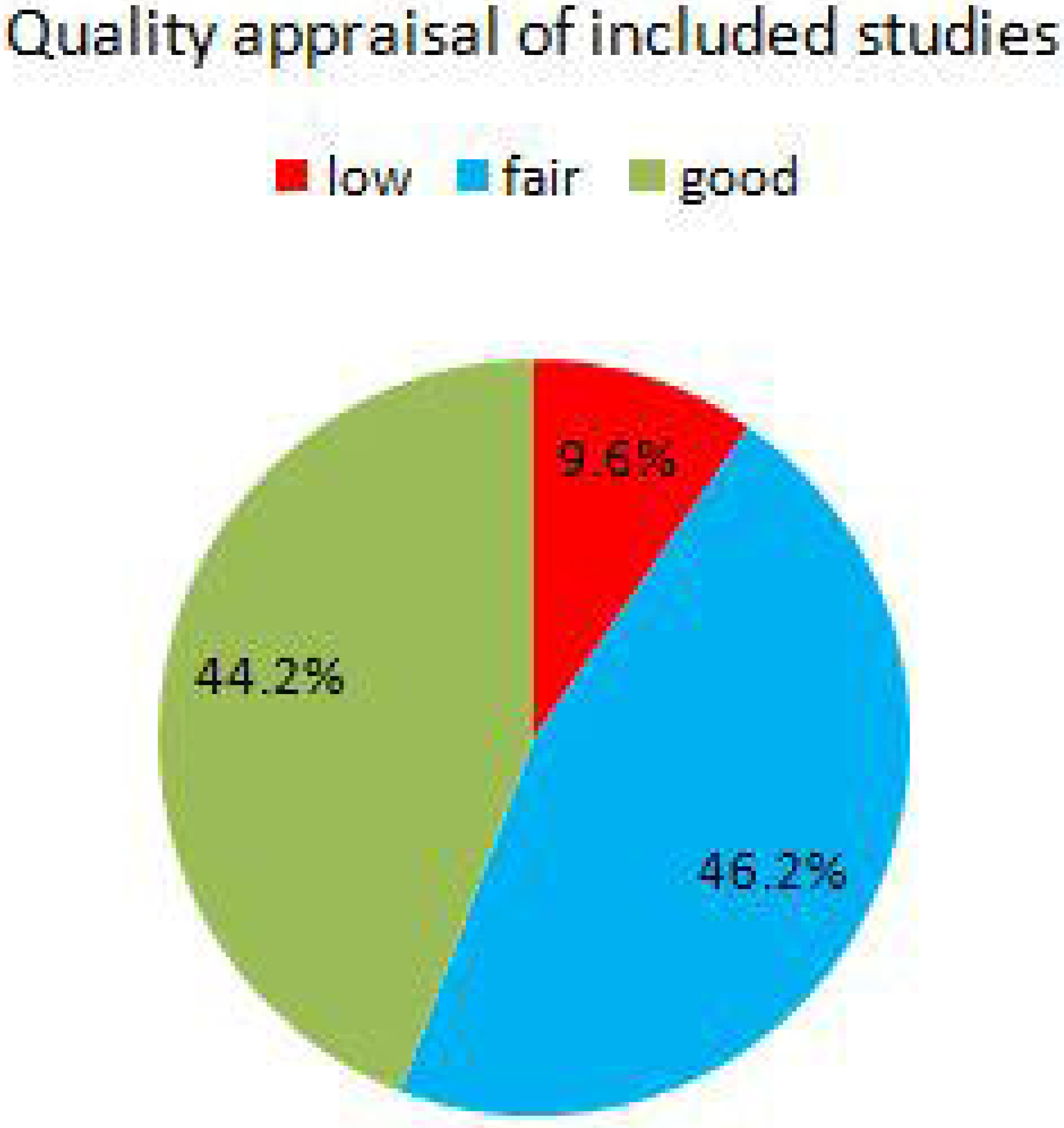
The percentage of included studies based on quality score.

### Synthesis of results

Among the included studies, 30.76% examined all three components of the KAPs model simultaneously. The most studied component in the studies was the knowledge component with about 84.61%, followed by attitude and practice with 53.84% and 44.23%, respectively. Also, 34.61% and 17.3% of the articles have examined other components including perception and awareness, respectively (Table 2, Fig 4).

**Table 2:** Results related to Coronavirus-related KAPs components of the included studies.

| Reference (Author, Year) | KAPs components |  |  | Other components |  |
| --- | --- | --- | --- | --- | --- |
|  | Level of Knowledge | Level of Attitudes | Level of Practices | Level of Perception | Level of Awareness |
| Abdelhafiz et al, 2020 [23] | Good | Positive | NA* | Moderate | NA |
| Al Nsour et al, 2020 [24] | NA | NA | NA | NA | Good |
| Alhaj et al, 2020 [25] | Good | NA | NA | NA | NA |
| Khan et al, 2020 [46] | NA | NA | NA | High | NA |
| Alzoubi et al, 2020 [26] | Good | Positive | Good | NA | NA |
| Escalera-Antezana et al, 2020 [36] | Good | NA | NA | NA | NA |
| Apaijitt and Wiwanitkit 2020 [27] | Fair | NA | NA | NA | NA |
| Azlan et al, 2020 [28] | Good | Positive | Fair | NA | NA |
| Basheti et al, 2020 [29] | NA | NA | NA | Moderate | Fair |
| Bhagavathula et al, 2020 [30] | Poor | NA | NA | High | NA |
| Bragazzi et al, 2020 [31] | Fair | Fairly positive | NA | NA | NA |
| Cai et al, 2020 [32] | NA | NA | Good | High | NA |
| Chesser et al, 2020 [33] | Fair | NA | NA | NA | NA |
| Clements, 2020 [34] | Good | NA | Fair | NA | NA |
| Galle et al, 2020 [37] | Good | NA | Fair | NA | Good |
| Geldsetzer et al, 2020 [38] | Good | NA | NA | Low | NA |
| Giao et al, 2020 [39] | Good | Positive | NA | NA | NA |
| Liu et al, 2020 [50] | Poor | NA | NA | Low | NA |
| Kamate et al, 2020 [42] | Good | Positive | Good | NA | NA |
| Karasneh et al, 2020 [43] | Fair | NA | NA | High | Fair |
| Kebede et al, 2020 [44] | Fair | NA | Fair | Moderate | NA |
| Erbas et al, 2020 [35] | Fair | Fairly positive | NA | NA | NA |
| Lau et al, 2020 [49] | Good | Positive | Good | NA | NA |
| Abdel Wahed et al, 2020 [70] | Good | Positive | NA | High | NA |
| Sari et al, 2020 [65] | Good | Positive | NA | NA | NA |
| Khasawneh et al, 2020 [47] | Good | Positive | Good | NA | NA |
| Khader et al, 2020 [45] | NA | Good | NA | Moderate | Good |
| Kumar et al, 2020 [48] | Poor | Positive | Fair | NA | NA |
| McFadde et al , 2020 [51] | NA | NA | NA | Low | NA |
| Modi et al, 2020 [52] | NA | NA | NA | NA | Fair |
| Moro et al, 2020 [53] | Fair | Positive | NA | NA | NA |
| Olapegba et al, 2020 [55] | Fair | NA | Fair | Moderate | NA |
| Olum et al,2020 [56] | Fair | Negative | Good | NA | NA |
| Parikh et al, 2020 [57] | Good | NA | NA | High | NA |
| Ricco et al, 2020 [60] | Fair | Negative | NA | NA | Fair |
| Roy et al, 2020 [61] | Fair | Positive | NA | NA | Good |
| Sahraian et al, 2020 [62] | Good | NA | NA | NA | NA |
| Salman et al, 2020 [63] | Fair | Positive | Fair | NA | NA |
| Saqlain et al, 2020 [64] | Good | Positive | Good | Moderate | NA |
| Shi et al, 2020 [66] | Good | Positive | NA | NA | NA |
| Singh et al, 2020 [67] | Good | NA | NA | High | NA |
| Siniscalchi et al, 2020 [68] | NA | NA | NA | Moderate | NA |
| Taghrir et al, 2020 [69] | Good | NA | Good | Moderate | NA |
| Nwafor et al, 2020 [54] | Fair | NA | Poor | NA | NA |
| Wolf et al, 2020 [71] | Fair | Fairly positive | Fair | NA | Fair |
| Yassa et al, 2020 [72] | Fair | Positive | NA | NA | NA |
| Zhang et al, 2020 [73] | Good | Positive | Fair | NA | NA |
| Zhong et al, 2020 [74] | Good | Positive | Good | NA | NA |
| Reuben et al, 2020 [59] | Good | Positive | Good | NA | NA |
| Jemal et al, 2020 [40] | Good | Positive | Fair | NA | NA |
| Raza et al, 2020 [58] | Good | Positive | Fair | NA | NA |
| Kakemam et al, 2020 [41] | Good | Positive | Fair | NA | NA |

\*NA: not assessed

**Figure 4:**
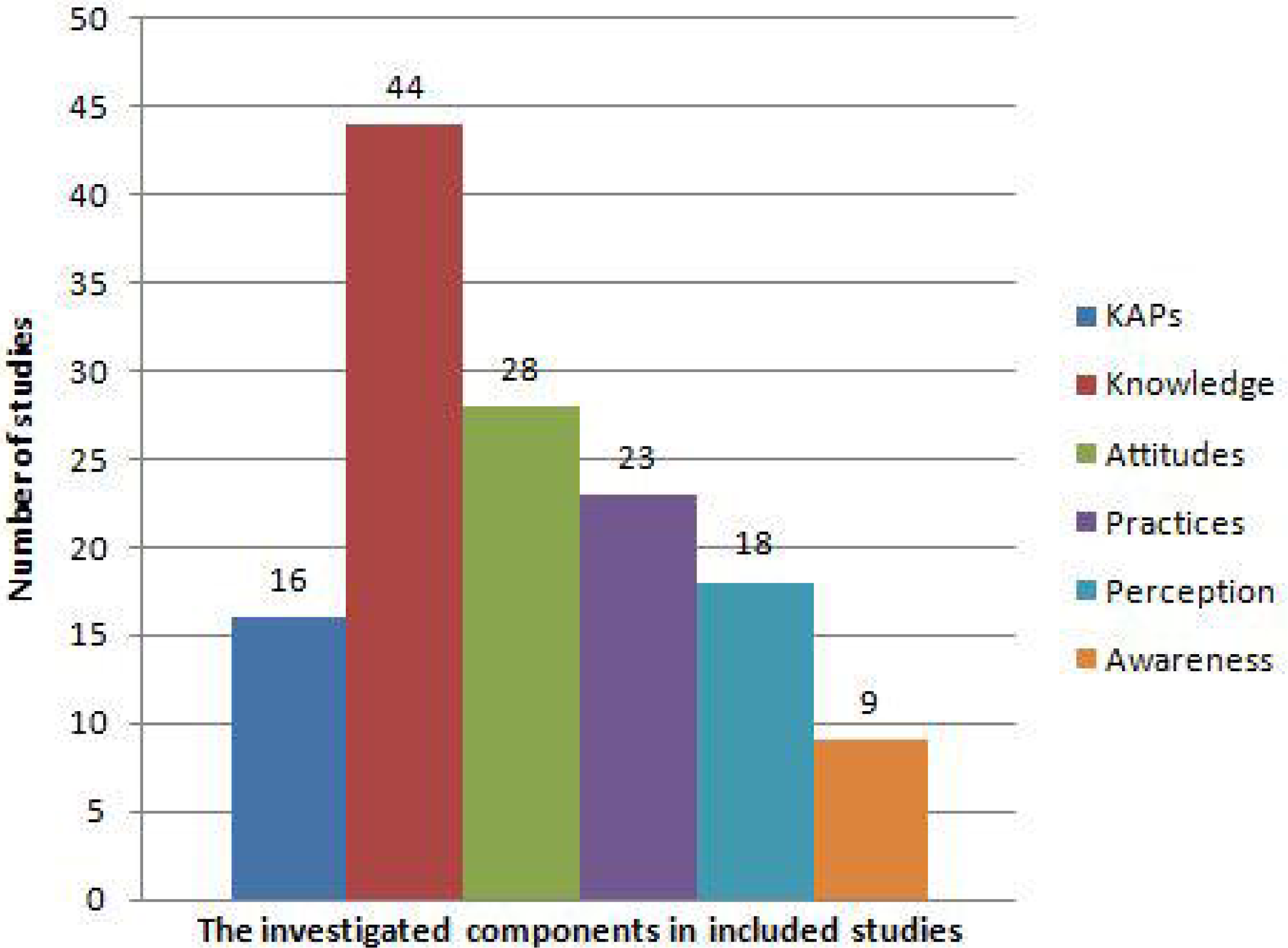
The number of investigated components in the included studies.

Of the studies that examined the knowledge component, 59% reported good knowledge, 34% fair knowledge, and 7% poor knowledge. As well as, of the studies that examined the attitude component, 82% reported a positive attitude, 11% a fairly positive attitude, and 7% a negative attitude. For the practice component, 44% reported good practice, 52% fair practice, and 4% poor practice. Also, 39% of the perception component reported a high level of perception and for the awareness component, 44% reported good awareness (Table 2, Fig 5).

**Figure 5:**
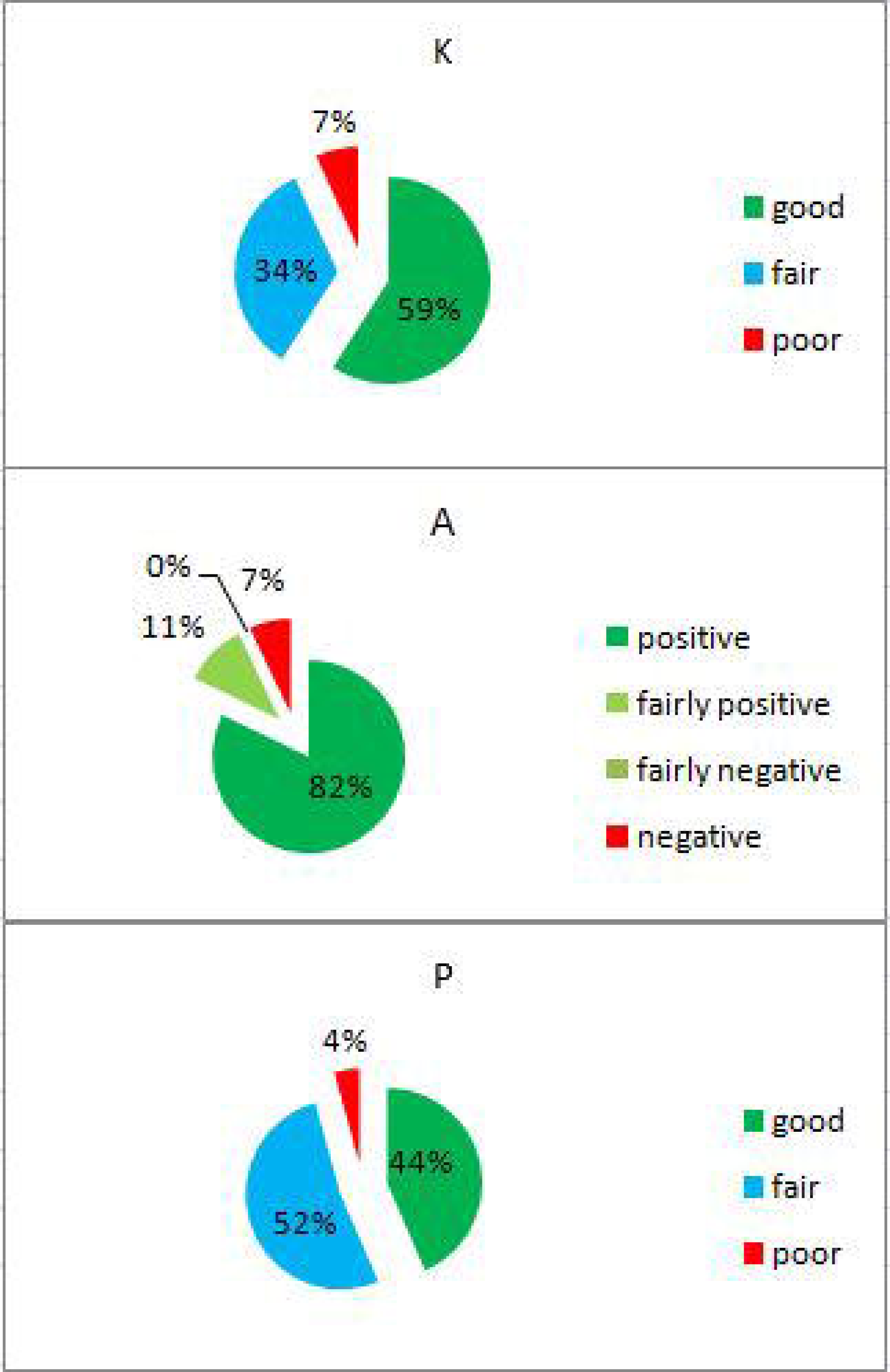
The percentage of studies based on the knowledge (K), attitudes (A), and practices (P)

## Discussion

COVID-19 has had serious, long-term, and sometimes irreparable effects on all aspects of the daily lives of individuals and society [75, 76]. Getting informed from the knowledge, attitude, and practice of different people can play a vital role in preventing diseases [77, 78], so the study of these components in different communities and between different groups seems necessary.

### Strength and weakness

Meta-analysis was not possible due to the different questionnaires and their dimensions, the target group, and study setting, so we reported the results descriptively in the form of tables and figures. Also, a large number of the included studies did not report the validity and reliability of the questionnaires, and therefore, the overall quality of the included studies was moderate. The main reason for this is the rush to publish articles related to coronavirus disease. Also, due to the high volume of articles published in this field, some new articles that may not have been included in this article may have been published during the writing and reviewing process. One of the most important strengths of this study was that all stages of the study were conducted with two researchers and in all stages, in cases of disagreement, the third person and consensus were used. Also, registering the protocol of this study and reviewing and modifying it in the PROSPERO platform is the strength of this study.

### Summary of study findings

We found that about 60% of the samples had good knowledge of COVID-19. Also, 82% of the samples were reported positive attitudes regarding COVID-19 and slightly more than 50% of samples performed good practices. About 80% of the studies used an online questionnaire to collect data, and the most used platforms included Google form, SurveyMonkey, and Qualtrics. The most important social media through which the questionnaires were distributed were Facebook, WhatsApp, and Telegram. The most important sources for learning and staying up to date about COVID-19 mentioned in the studies were television, social media, internet, radio, and friend and relatives.

The finding of our systematic review demonstrated fairly good knowledge about COVID-19. In most studies, more than 70% of the participants had a good knowledge of issues such as causes, symptoms, ways of transmission, and ways of prevention. Also, the majority of participants had a high level of knowledge about symptoms such as high fever and dry cough, breathing difficulty and a small number had sufficient knowledge about other symptoms such as chills, headache, muscle pain, sore throat, and loss of taste or smell [23, 25, 26, 34, 36-38, 41, 42, 64, 66]. More than 90% of the participants considered air droplets as a way to spread. This good level of knowledge can be due to widespread information through various means such as public media (television and radio), social media, and government announcements. In addition, preparing several guidelines and reports by WHO, CDC and local government in times of outbreak and easy access to them have increased the level of information and knowledge of individuals regarding COVID-19 [79, 80]. On the other hand, factors such as low literacy level, older age, and the presence of the rural population in the samples were among the factors that have reduced the level of knowledge in the studies [23, 27, 31, 33, 71].

In this review, participants showed a positive attitude regarding COVID-19. Almost all participants believed in the importance of handwashing, disinfecting surfaces, using masks to prevent the spread of infection, resting at home in the event of symptoms, and maintaining social distance and limited contact. Of course, in some cases, there was a negative belief that it could be due to differences in instructions and guidelines by different institutions, such as what was about wearing a face mask at the beginning of the pandemic, and then it was recommended that the whole population should use a mask [56, 60]. Such cases show the importance of integrated guidelines and the focus of decision-making in times of crisis [81, 82]. Although having a responsible organization can help make better and faster decisions, in such cases, political pressure is exerted by governments that such organizations should put the health of the people at the top and not refuse to make the right decisions due to political pressures [11, 83, 84].

In general, the level of practice of the participants in the studies was fair. However, despite the high knowledge and positive attitude of the participants, the level of practices was still sometimes lower than expected. Numerous reasons for a low level of practice have been cited in studies. Lack of availability (for example, masks and disinfectants), imposing financial costs on participants, ambiguity in instructions, not getting used to new conditions such as staying home and wearing a mask, exhaustion from existing conditions, and anxiety and stress of disease were among the causes mentioned in the studies [28, 34, 40, 44, 54, 58, 71]. In this regard, some countries have imposed strict laws and penalties on people who do not follow the guidelines to improve their performance, but in many countries under study, such laws do not exist and have not been applied [85, 86]. Another factor that affects the performance of individuals was the presence of decision-makers in public and social media. Seeing a person without a mask at the height of a pandemic can have a negative impact on a person’s good practices.

Given the diversity of settings and questionnaires, the authors of this paper recommend that there be a need to design an integrated online system to assess the knowledge, attitudes, and practices of the population about health-related crises. Designing such an integrated system can help better compare countries because integrated items are used for comparison. On the other hand, designing such a system and disseminating its results can accelerate integrated decision-making and improve crisis management. On the other hand, the existence of such an integrated system can lead to an increase in solidarity, which was emphasized by the World Health Organization during the corona pandemic [87, 88].

### Limitations

The included studies were from both high and low-income countries and therefore generalization of results to all countries should be done with caution. Also, many of the questionnaires in the studies did not have sufficient validity and reliability or did not report it. Our review study also had some limitations. Due to differences in studies and the use of different questionnaires, conducting a meta-analysis was not possible in this study. We have only reviewed studies published in English. On the other hand, due to the high speed of publication of articles in this field, some other studies may be published at the time of writing the article and the review process, which has been missed. Of course, due to the high speed of publishing articles, this limitation is inevitable.

## Conclusion

This systematic review showed that the KAP components in the participants are at an acceptable level. In general, knowledge was at a good level, the attitude was positive and practice was at a fairly good level. Providing accurate and up-to-date information in times of crises and disseminating them through responsible institutions and through the mass media and holding online training courses can help increase people’s knowledge, attitudes, and practices.

## Data Availability

All relevant data exist in the manuscript.

## Supporting Information

Supplementary file 1; PubMed search strategy

Supplementary file 2; quality appraisal of the included studies

## Acknowledgments

We thank the PROSPERO institute for accelerating the review process in the time of Coronavirus. We also thank Birjand University of Medical Sciences for approving our proposal and giving it a code of ethics (IR.BUMS.REC.1399.099).

## Author Contributions

Conception and design: MA-Z. Screen the records, data extraction, and quality appraisal: MM, SS and SH. Data analysis: MA-Z and HA. Draft manuscript: MA-Z. Critical review: SH and HA. All authors approved the final version of the manuscript for publication.

## Supporting information captions

Supplementary file 1: PRISMA 2009 checklist

Supplementary file 2: Complete search strategy

Supplementary file 3: Quality appraisal of the included studies

